# Impact of Clinical Frailty Scale on Clinical Outcomes and Decision-Making to Prescribe Anticoagulation Following LAAC

**DOI:** 10.64898/2025.12.19.25342722

**Authors:** Koya Okabe, Mike Saji, Mamoru Nanasato, Mai Terada, Yuki Izumi, Mitsunobu Kitamura, Iraru Takamisawa, Mitsuaki Isobe, Masahiko Asami, Mitsuru Sago, Shuhei Tanaka, Ryuki Chatani, Toru Naganuma, Yohei Ohno, Tomoyuki Tani, Hideharu Okamatsu, Gaku Nakazawa, Yusuke Watanabe, Masaki Izumo, Shingo Mizuno, Daisuke Hachinohe, Hiroshi Ueno, Shunsuke Kubo, Shinichi Shirai, Masaki Nakashima, Masanori Yamamoto, Kentaro Hayashida, the OCEAN-LAAC investigators

## Abstract

**Background:** Anticoagulants are often less frequently prescribed in elderly patients with atrial fibrillation (AF) because of concerns regarding high bleeding risk, despite their increased susceptibility to embolic stroke and systemic embolization. This study evaluated the impact of the Clinical Frailty Scale (CFS) on clinical outcomes and decision-making for prescribing antithrombotic therapy following left atrial appendage closure (LAAC) in a large contemporary registry.

**Methods:** The OCEAN-LAAC registry included 1,409 patients who underwent LAAC. Outcomes and antithrombotic prescriptions after the procedure were compared between groups stratified by CFS into 1–3 and 4–8.

**Results:** Patients with CFS 4–8 were more likely to have a history of stroke and demonstrated lower serum albumin and hemoglobin levels, consistent with advanced frailty. In multivariate analysis, CFS 4–8 was independently associated with higher all-cause mortality at one year compared with CFS 1–3 (adjusted hazard ratio 1.89; 95% confidence interval 1.05–3.41). By one year, patients with CFS 4–8 more frequently discontinued antithrombotic therapy, without significant differences in ischemic stroke or device-related thrombotic events. Notably, major bleeding was more common in the CFS 4–8 group, reflecting their advanced clinical vulnerability.

**Conclusion:** Greater frailty, as assessed by CFS, was independently associated with increased all-cause mortality following LAAC. Although frailty influenced patterns of antithrombotic therapy in this real-world registry, thrombotic events remained comparable between CFS groups, supporting the feasibility of individualized, frailty-guided post-LAAC management. These findings underscore the importance of incorporating frailty assessment into multidisciplinary Brain–Heart team decision-making.

## 1. Introduction

Frailty is a multidimensional geriatric syndrome characterized by decreased physiological reserves and increased vulnerability to stressors (1). It is associated with poor prognosis (2,3), and influences the management of anticoagulant prescriptions in elderly patients with atrial fibrillation (AF) (4–7). This is partly due to the controversy surrounding the high bleeding risk in this population, despite their increased susceptibility to embolic stroke and critical systemic embolization (7–10). The Clinical Frailty Scale (CFS) was developed from the Canadian Study of Health and Aging a decade ago, and is a well-validated, semi-quantitative tool to assess frailty in elderly populations (11,12).

Left atrial appendage closure (LAAC) has recently emerged as an alternative strategy for stroke prevention in patients with non-valvular AF who are at high risk of bleeding, and it may specifically benefit patients with frailty as it helps reduce antithrombotic medication safely following the procedure (13,14). Recent studies have indicated that frailty correlates with worse outcomes following LAAC (15–17). However, whether frailty status can affect the decision to prescribe anticoagulation after LAAC remains unclear. To address this issue, we examined a large registry dataset. Although prior studies reported that frailty is associated with worse clinical outcomes after LAAC, little is known about how frailty influences real-world antithrombotic management following the procedure. Because the optimal antithrombotic regimen after LAAC remains controversial, understanding the interaction between frailty and post-procedural antithrombotic decisions is clinically crucial. Therefore, our aim was to assess the impact of the CFS on clinical outcomes (e.g. all-cause mortality, bleeding, and thrombotic events), and on the decision-making process for antithrombotic therapy following LAAC in elderly patients with AF.

## 2. Methods

### 2.1. Study population and design

The OCEAN (Optimized CathEter vAlvular iNtervention)-LAAC registry is an ongoing, prospective, investigator-initiated, multicenter, observational registry. It enrolls patients with non-valvular AF undergoing percutaneous LAAC using either WATCHMAN 2.5^®^ or WATCHMAN FLX^®^ devices in Japan. Nineteen centers have participated in this registry under the University Hospital Medical Information Network (UMIN000038498), which commenced in September 2019. Baseline characteristics, including risk scores for frailty (CFS), bleeding (HAS-BLED score), and thromboembolism (CHA_2_DS_2_-VASc scores), were recorded. Patients were qualified for LAAC if they had non-valvular AF, required anticoagulation, and had a high bleeding risk that met at least one of the following conditions: 1) a HAS-BLED score of three or higher, 2) multiple instances of trauma due to falls, 3) a history of diffuse cerebral amyloid angiopathy, 4) long-term use of anticoagulation combined with dual antiplatelet therapy (or triple therapy), or 5) a history of major bleeding of BARC type 3 or higher. The exclusion criteria for this analysis included: 1) thrombus in the atrium or appendage which is considered inappropriate for LAAC procedure by the brain-heart team, 2) contraindications to transesophageal echocardiography (TEE) insertion or catheterization due to anatomical reasons, and 3) contraindications for taking anticoagulation or antiplatelet agents. Procedural-related data, postprocedural clinical outcomes, and detailed prescription data pre- and post-LAAC were collected in the registry. Data were prospectively obtained using a dedicated web-based database (Canon Medical Systems, Nasu, Japan).

At each center, the local multidisciplinary brain-heart team, comprising cardiac surgeons, interventional cardiologists, neurologists, neurosurgeons, and imaging specialists, determined the eligibility of LAAC and the treatment strategy. LAAC was performed using standard techniques, as previously described (18,19). This study was approved by the Ethics Committee of the Keio University School of Medicine (approval number: #20190194) as well as by the ethics committee of the participating organization. The information about the study was explained orally or disclosed on the institutional website, giving potential participants the opportunity to opt out. All procedures were conducted in accordance with the principles of the Declaration of Helsinki, and informed consent was obtained from each patient. The OCEAN-SHD registry is independent of industrial influence, is registered with the University Hospital Medical Information Network (UMIN0000 38498).

The CFS was scored from 1 to 9 based on a semi-quantitative evaluation of the patient’s symptoms, mobility, inactivity, exhaustion, and disability for basic activities of daily living and instrumental activities of daily living (12). Baseline characteristics and outcomes were compared by dividing patients into two groups according to CFS (1 to 3, 4 to 8). In the present study, no patients were graded as CFS 9.

### 2.2. Pre-procedural imaging assessment

All patients underwent transthoracic echocardiography (TTE) and/or TEE prior to LAAC. A minimum of three consecutive heartbeats were recorded, and the data were averaged for the analysis of echocardiographic variables. As per the guidelines of the American Society of Echocardiography and the European Association of Cardiovascular Imaging, both two-dimensional and three-dimensional echocardiographic assessments were conducted (20). Baseline TEE measurements included the ostium diameter and depth of the left atrial appendage at angles of 0°, 45°, 90°, and 135° (21).

### 2.3. Data Collection for Mandated Clinical Follow-Up

Clinical follow-up was conducted within 60 days following LAAC using standardized interviews, documentation from referring physicians, and hospital discharge summaries. Collected demographic data included laboratory data, and imaging assessments such as TTE and/or TEE. In-hospital and follow-up data encompassed all suspected serious adverse events including all-cause mortality, major bleeding, ischemic stroke, procedure-related complications (e.g. pericardial effusion requiring drainage, peri-procedural bleeding requiring surgical revision, transfusion, and device embolization) and device related thrombus. Outcomes were independently adjudicated by the local clinical events committees of each center, in accordance with previous literature (22).

### 2.4. Statistical Analysis

Continuous variables were tested for normal distribution using the Shapiro–Wilk test and are presented as either mean ± standard deviation or median with interquartile range, as appropriate. Categorical variables are expressed as counts and percentages. Comparison of continuous variables was performed using the unpaired Student t-test or the Wilcoxon rank sum test, based on the distribution of the variables. The chi-square test or Fisher’s exact test was utilized for comparing categorical variables. Kaplan–Meier analysis was used to estimate cumulative mortality by using the log-rank test. To investigate the association between CFS and outcomes, variables with p-value < 0.05 after univariate analysis (Table 1) were entered into multivariate Cox regression analysis. A forest plot was generated to evaluate the association between CFS 4–8 and all-cause mortality across predefined subgroups of patients with LAAC. All analyses were conducted using SPSS software (IBM SPSS Statistics, version 18, Armonk, NY, USA).

**Table 1.**
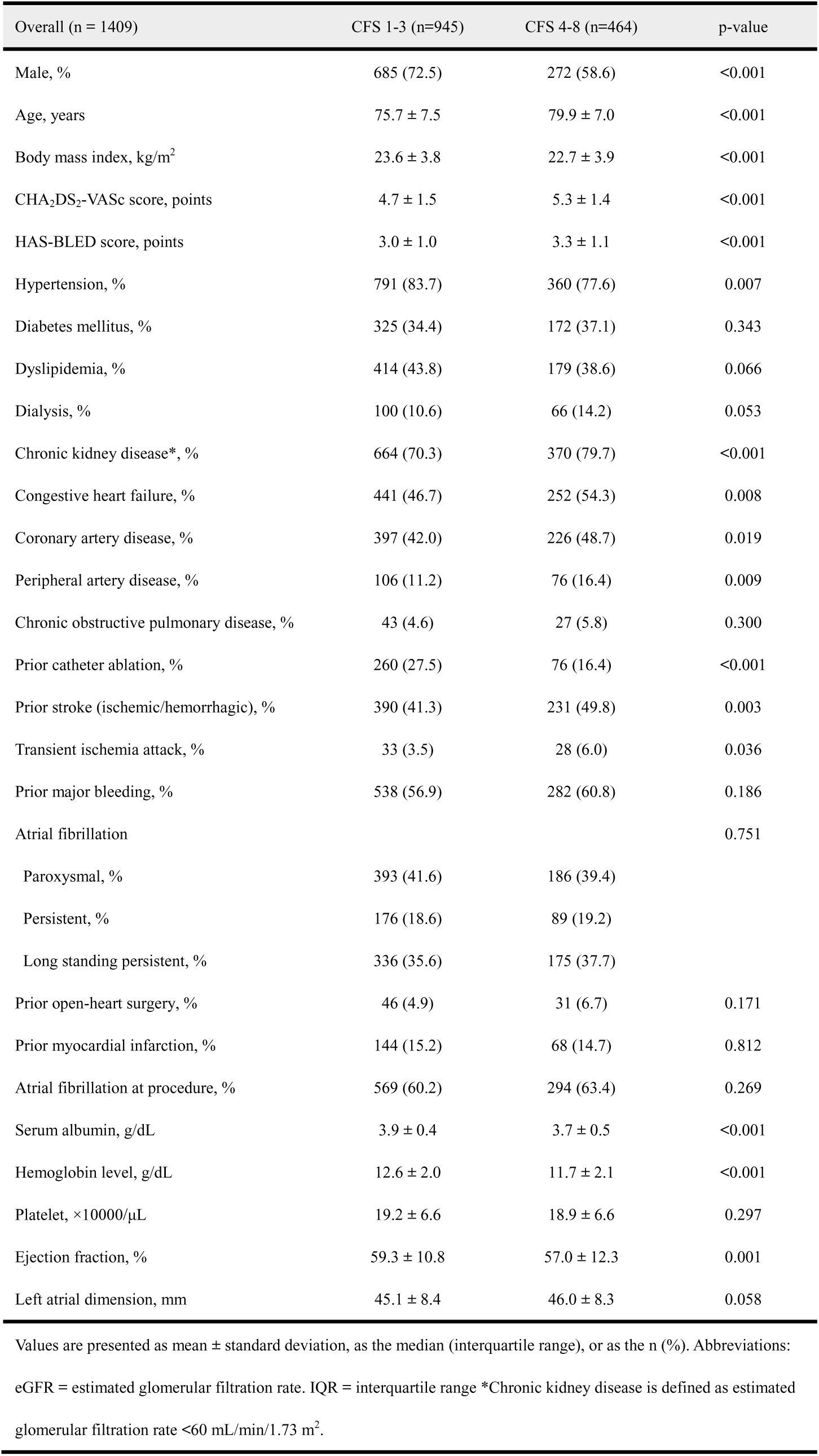
Baseline Patient Characteristics.

## 3. Results

### 3.1. Baseline characteristics

Of 1,464 patients undergoing LAAC, 55 patients were excluded due to lack of information about CFS, and 1,409 patients were finally analyzed. With median age of 78 years, 945 (67.1%) were categorized as CFS 1–3, 464 (32.9%) were as CFS 4–8 **(Table 1)**. Patients with greater CFS were more likely to be older and female, have hypertension, chronic kidney disease, prior stroke and peripheral artery disease with higher CHA_2_D_2_-VASc and HAS-BLED score. In addition, they had lower body mass index, lower serum albumin, and lower hemoglobin level.

### 3.2. Clinical outcomes at one year after LAAC

As shown in **Figure 1**, there were significant differences between groups in all-cause mortality and major bleeding events following LAAC in Kaplan-Meier analysis. Conversely, no significant differences between groups were observed in terms of ischemic stroke, and device related thrombus.

**Figure 1.**
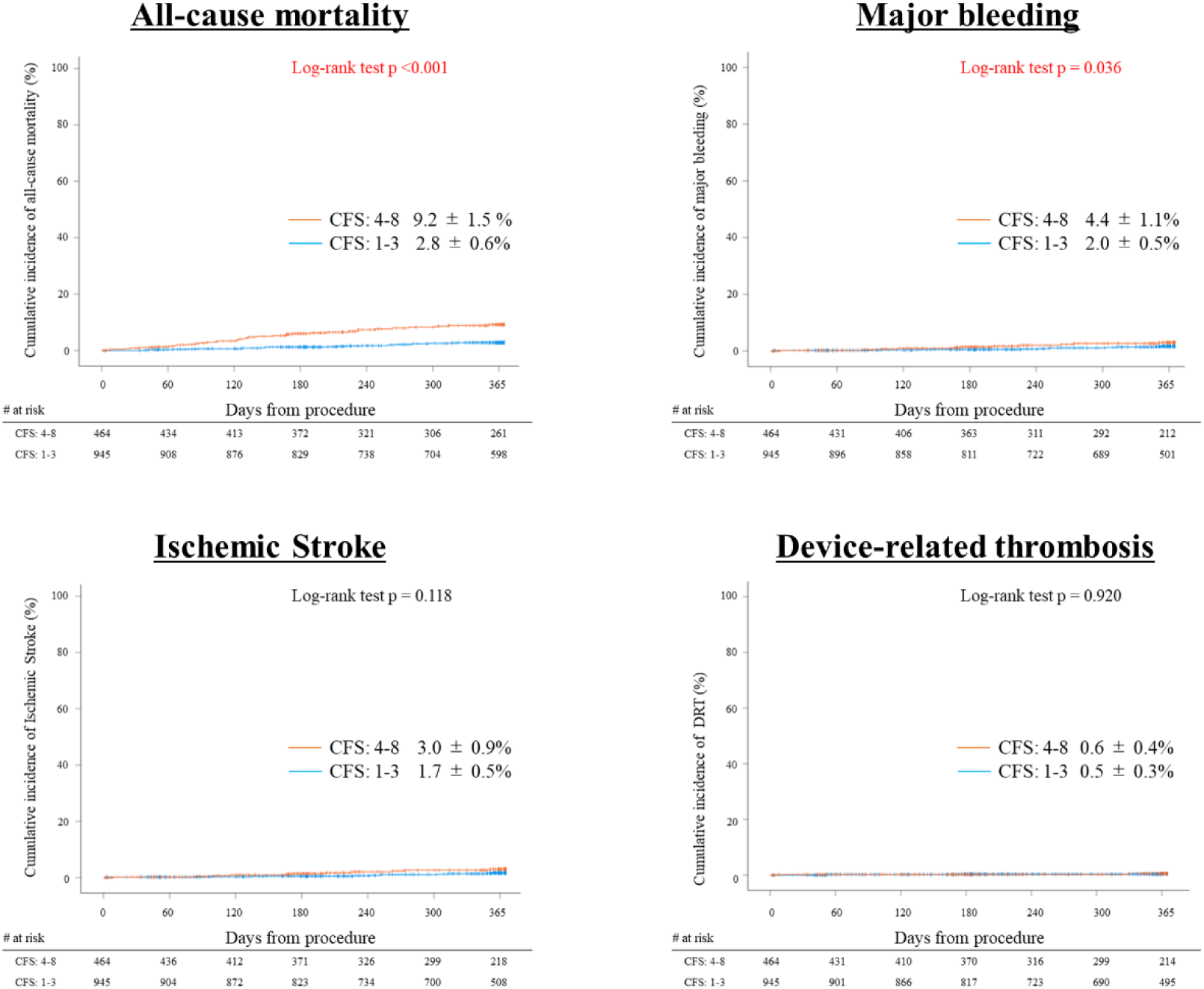
Kaplan-Meier analysis for all-cause mortality following LAAC according to tertiles stratified by clinical frailty scale. CFS 4–8 had higher all-cause mortality and major bleeding rate than those with CFS 1–3 (Log-rank p <0.001, and 0.036, respectively), while they had similar ischemic stroke and device-related thrombus rates compared with CFS 1–3 following LAAC (Logrank p 0.118, and 0.920, respectively). CFS = clinical frailty scale

### 3.3. Cox regression analysis for all-cause mortality at one year after LAAC

After performing univariate analysis, CFS groups as main variables, and age, body mass index <20 kg/m^2^, CHA2DS2-VASc score, dialysis, congestive heart failure, peripheral artery disease, chronic obstructive pulmonary disease, serum albumin, hemoglobin level, and ejection fraction as adjusting variables, were included in a multivariate Cox regression analysis **(Table 2)**. The analysis revealed that CFS 4-8 is independently associated with increased risk of all-cause mortality at one year after LAAC (adjusted hazard ratio 1.89, 95% CI 1.05–3.41) compared with CFS 1–3 as a reference even after adjustment. The forest plot demonstrated that CFS 4–8 were associated with all-cause mortality across the majority of subgroups **(Figure 2)**.

**Figure 2.**
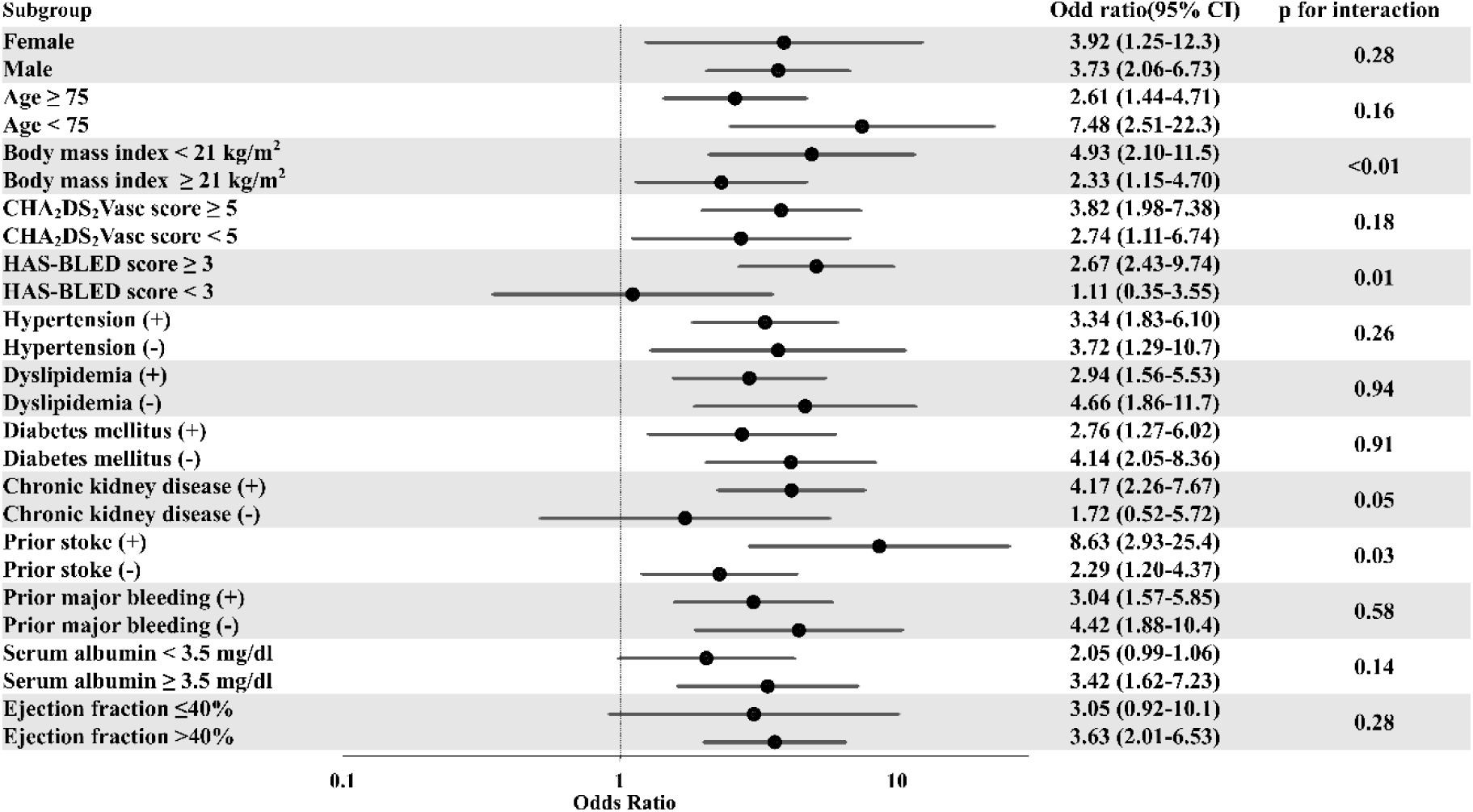
Forest plot of CFS 4–8 and all-cause mortality across predefined subgroups. The forest plot demonstrates the association between CFS 4–8 and all-cause mortality across clinically relevant subgroups. The association between CFS 4–8 and increased mortality was consistent across the majority of subgroups analyzed. CI = confidence interval

**Table 2.** Univariate and Multivariate Cox Regression Analysis for Predicting All-cause Mortality after Left Atrial Appendage Closure.

|  | Univariate analysis |  |  | Multivariate analysis |  |  |
| --- | --- | --- | --- | --- | --- | --- |
|  | Unadjusted<br>Hazard ratio | 95% CI | p-value | Adjusted<br>Hazard ratio | 95% CI | p-value |
| Clinical frailty scale 1-3 | 1.00 | - | - |  | - | - |
| Clinical frailty scale 4-8 | 3.51 | 2.09-5.91 | <0.001 | 1.89 | 1.05-3.41 | **0.034** |
| Adjusting parameters |  |  |  |  |  |  |
| Male | 0.72 | 0.40-1.29 | 0.270 |  |  |  |
| Age (every 10 years increase) | 1.49 | 1.05-2.12 | 0.029 | 1.35 | 0.93-1.96 | 0.111 |
| Body mass index (<20 kg/m<sup>2</sup>) | 2.47 | 1.46-4.18 | <0.001 | 1.37 | 0.78-2.41 | 0.270 |
| CHA<sub>2</sub>DS<sub>2</sub>-VASc score (every degree increase) | 1.19 | 1.01-1.41 | 0.038 | 1.01 | 0.82-1.24 | 0.926 |
| HAS-BLED score (every degree increase) | 1.13 | 0.89-1.43 | 0.311 |  |  |  |
| Hypertension | 0.61 | 0.34-1.08 | 0.087 |  |  |  |
| Diabetes mellitus | 1.40 | 0.84-2.33 | 0.201 |  |  |  |
| Dyslipidemia | 0.66 | 0.39-1.13 | 0.128 |  |  |  |
| Dialysis | 2.35 | 1.29-4.27 | 0.005 | 1.01 | 0.78-2.41 | 0.195 |
| Chronic kidney disease* | 1.47 | 0.78-2.76 | 0.239 |  |  |  |
| Congestive heart failure | 2.83 | 1.60-5.02 | <0.001 | 1.54 | 0.79-3.00 | 0.202 |
| Coronary artery disease | 1.62 | 0.97-2.70 | 0.063 |  |  |  |
| Peripheral artery disease | 2.57 | 1.45-4.56 | 0.001 | 1.82 | 0.97-3.41 | 0.063 |
| Chronic obstructive pulmonary disease | 2.34 | 1.01-5.45 | 0.048 | 2.46 | 1.04-5.82 | **0.040** |
| Prior catheter ablation | 0.56 | 0.27-1.13 | 0.105 |  |  |  |
| Prior ischemic/hemorrhagic stroke | 1.09 | 0.65-1.81 | 0.752 |  |  |  |
| Transient ischemia attack | 0.72 | 0.18-2.96 | 0.652 |  |  |  |
| Prior major bleeding | 1.14 | 0.68-1.92 | 0.619 |  |  |  |
| Prior open-heart surgery | 0.579 | 0.14-2.37 | 0.448 |  |  |  |
| Prior myocardial infarction | 1.35 | 0.72-2.54 | 0.352 |  |  |  |
| Atrial fibrillation at procedure | 0.89 | 0.54-1.49 | 0.670 |  |  |  |
| Serum albumin (g/dL) | 0.18 | 0.12-0.28 | <0.001 | 0.33 | 0.19-0.55 | **<0.001** |
| Hemoglobin level (g/dL) | 0.79 | 0.69-0.90 | <0.001 | 1.03 | 0.89-1.19 | 0.672 |
| Platelet (×1000/μL) | 1.01 | 0.98-1.05 | 0.529 |  |  |  |
| Ejection fraction (%) | 0.95 | 0.94-0.97 | <0.001 | 0.97 | 0.95-0.99 | **0.003** |
| Left atrial dimension (mm) | 1.02 | 0.99-1.05 | 0.202 |  |  |  |

CI = confidence interval, eGFR = estimated glomerular filtration rate. \*Chronic kidney disease was defined as eGFR <60 mL/min/1.73 m<sup>2</sup>.

### 3.4. Trends in antithrombotic therapy

Antithrombotic therapy at each period is shown in **Table 3**. Overall, there were no significant differences in antithrombotic therapy at baseline, at discharge and at 3 months after LAAC between two groups. However, at one year, in CFS 4-8 group compared with CFS 1–3 group, more patients de-escalated antithrombotic drugs. In particular, anticoagulants were less frequently continued in the CFS 4–8 at one year than CFS 1–3 groups.

**Table 3.** Antithrombotic Therapy Stratified by Clinical Frailty Scale.

| Table 3. Antithrombotic Therapy Stratified by Clinical Frailty Scale |  |  |  |
| --- | --- | --- | --- |
| Antithrombotic drug | CFS 1–3 | CFS 4–8 | p-value |
| At baseline | n=929 | n=455 |  |
| OAC+DAPT | 17 (1.8) | 7 (1.5) | 0.518 |
| OAC+SAPT | 289 (31.1) | 144 (31.0) |  |
| DAPT | 8 (0.9) | 7 (1.5) |  |
| OAC alone | 594 (63.9) | 283 (61.0) |  |
| SAPT | 15 (1.6) | 7 (1.5) |  |
| No antithrombotic drug | 6 (0.6) | 7 (1.5) |  |
| On OAC | 900 (95.2) | 434 (93.5) | 0.162 |
| At discharge | n=934 | n=459 |  |
| OAC+DAPT | 15 (1.6) | 6 (1.3) | 0.327 |
| OAC+SAPT | 537 (57.5) | 252 (54.9) |  |
| DAPT | 15 (1.6) | 11 (2.4) |  |
| OAC alone | 357 (38.2) | 181 (39.4) |  |
| SAPT | 6 (0.6) | 8 (1.7) |  |
| No antithrombotic drug | 4 (0.4) | 1 (0.2) |  |
| On OAC | 909 (97.3) | 439 (95.6) | 0.095 |
| At 3 months after LAAC | n=910 | n=435 |  |
| OAC+DAPT | 3 (0.3) | 1 (0.2) | 0.066 |
| OAC+SAPT | 97 (10.7) | 53 (12.2) |  |
| DAPT | 352 (38.7) | 141 (32.4) |  |
| OAC alone | 179 (19.7) | 76 (17.5) |  |
| SAPT | 254 (27.9) | 144 (33.1) |  |
| No antithrombotic drug | 25 (2.7) | 20 (4.6) |  |
| On OAC | 279 (30.7) | 130 (29.9) | 0.800 |
| At 1 year after LAAC | n=687 | n=298 |  |
| OAC+DAPT | 0 (0) | 0 (0) | 0.009 |
| OAC+SAPT | 22 (3.2) | 8 (2.7) |  |
| DAPT | 32 (4.7) | 12 (4.0) |  |
| OAC alone | 84 (12.2) | 26 (8.7) |  |
| SAPT | 503 (73.2) | 212 (71.1) |  |
| No antithrombotic drug | 46 (6.7) | 40 (13.4) |  |
| On OAC | 106 (15.4) | 34 (11.4) | 0.001 |
| Values are presented as the n (%). Abbreviations: CFS = Clinical Frailty Scale, DAPT = dual antiplatelet therapy, LAAC = left atrial |  |  |  |

appendage closure, OAC = Oral anticoagulant, SAPT = single antiplatelet therapy.

## 4. Discussion

In this large, real-world registry of elderly patients undergoing LAAC, we demonstrated that frailty assessed by the Clinical Frailty Scale (CFS) has a substantial impact on both clinical outcomes and post-procedural antithrombotic management. The principal findings of this study are as follows: (1) patients with greater frailty burden exhibited characteristic clinical features—older age, nutritional depletion, and prior bleeding or thromboembolic events—consistent with the pathophysiology of frailty; (2) advanced frailty was associated with significantly increased all-cause mortality and major bleeding within one year after LAAC; (3) CFS remained an independent predictor of one-year all-cause mortality even after robust adjustment for comorbidities and laboratory parameters; and (4) despite similar rates of ischemic stroke and device-related thrombus across frailty strata, frail patients more frequently underwent de-escalation of antithrombotic therapy by one year following LAAC.

### 4.1 Frailty as a determinant of prognosis after LAAC

Our findings reinforce and extend prior reports that frailty is associated with adverse outcomes following LAAC (15). Notably, while previous studies primarily utilized the Hospital Frailty Risk Score (HFRS), a claims-based tool validated only in the very elderly, we employed the CFS—a clinically intuitive, widely adopted scale that reflects functional capacity and global physiological reserve. Using CFS allowed more accurate characterization of frailty status in a real-world elderly AF population and provides meaningful granularity for bedside decision-making. Consequently, our results offer stronger evidence that frailty itself, independent of conventional risk factors, contributes to mortality risk after LAAC.

### 4.2. Preserved procedural safety in frailty

Interestingly, procedural complications were comparable across frailty categories. These results contrast with transcatheter aortic valve replacement populations, where frailty consistently predicts peri-procedural morbidity (3). The absence of excess procedural risk suggests that LAAC is sufficiently low-risk to be safely performed in frail individuals and should not be withheld solely based on frailty status.

### 4.3. Bleeding vulnerability and implications for antithrombotic decision-making

A key contribution of this study is the demonstration that frailty substantially influences real-world antithrombotic prescribing patterns after LAAC. Despite similar antithrombotic use at discharge and three months, frail patients were more likely to discontinue or attenuate antithrombotic therapy by one year. This pattern appears clinically coherent: frail patients experienced a higher burden of major bleeding, whereas thrombotic events did not increase. The observed data indicate that clinicians may already be individualizing therapy on the basis of frailty, particularly as bleeding events accumulate over time.

Crucially, our findings suggest that current standardized antithrombotic protocols after LAAC may not adequately reflect the heterogeneous bleeding risk imposed by frailty. Because LAAC effectively mitigates thromboembolic risk, frail patients—with an inherently high bleeding nature—may benefit from more tailored and possibly earlier de-escalation strategies (4–6,9). These insights highlight the need for prospective trials evaluating frailty-guided antithrombotic regimens after LAAC.

### 4.4. Clinical implications: frailty as a foundation for personalized LAAC management

Taken together, our findings emphasize that frailty should be incorporated into clinical workflows when considering LAAC. Frail individuals may stand to benefit the most from LAAC, given their poor tolerance for long-term anticoagulation and their preserved protection against thromboembolic events after the procedure. However, their increased bleeding risk and higher all-cause mortality underscore the importance of careful post-procedural surveillance, attention to nutritional and functional status, and adaptive antithrombotic management. A multidisciplinary Brain–Heart team approach may be particularly valuable for this population, integrating cardiologists, neurologists, geriatricians, and rehabilitation specialists to balance bleeding and thrombotic risk and to optimize functional outcomes. Incorporating frailty as a routine parameter can facilitate precision medicine, ultimately improving the safety and effectiveness of LAAC in elderly patients with AF.

## 5. Limitations

This study has several limitations. First, it was conducted using a single-country registry in Japan and employed a non-randomized, unblinded, observational design. These factors may introduce inherent biases, including population-specific characteristics such as smaller body size, distinct comorbidity profiles, and potentially longer life expectancy compared with Western cohorts. Therefore, validation in multinational datasets or randomized controlled studies is warranted. Second, although the multivariate model incorporated key clinical variables, residual confounding cannot be excluded. Unmeasured factors—such as frailty-related functional deficits, cognitive status, or socioenvironmental determinants—may have influenced mortality risk and antithrombotic prescribing patterns. Thus, the association between advanced frailty and post-LAAC antithrombotic decisions should be interpreted cautiously. Nevertheless, the large sample size and real-world nature of the registry enhance the reliability of our observations. Third, frailty assessment was limited to the Clinical Frailty Scale. Other frailty domains, including physical performance measures, sarcopenia indicators, or comprehensive geriatric assessments, were not available. Because each frailty construct captures different physiological vulnerabilities, future studies should integrate multiple frailty markers to more fully evaluate their prognostic and therapeutic implications after LAAC. Finally, follow-up was limited to one year. Longer-term data are needed to determine whether the prognostic impact of frailty and its influence on antithrombotic management persist beyond the first year after LAAC.

## 6. Conclusion

This study provides novel evidence that frailty, assessed by CFS, not only predicts mortality after LAAC but also shapes real-world antithrombotic therapy decisions. The preserved thrombotic protection despite greater de-escalation in frail individuals indicates that LAAC may be particularly advantageous for this vulnerable population. Future studies should explore frailty-guided antithrombotic strategies to optimize post-LAAC care.

## Data Availability

Due to privacy or ethical restrictions, the data cannot be publicly shared, but are available from the corresponding author on reasonable request.

## Acknowledgements

The authors thank all the investigators and institutions that have been involved in the OCEAN-LAAC registry.

## Clinical trials

OCEAN-LAAC registry (UMIN-ID: UMIN000038498)

## Abbreviations

AF: atrial fibrillation
CFS: Clinical frailty Scale
LAAC: left atrial appendage closure
TEE: transesophageal echocardiography
TTE: transthoracic echocardiography

## References

1. Fried LP, Tangen CM, Walston J, Newman AB, Hirsch C, Gottdiener J, et al. Frailty in older adults: evidence for a phenotype. J Gerontol A Biol Sci Med Sci. 2001;56(3):M146–56.

2. Tokuda T, Yamamoto M, Kagase A, Shimura T, Yamaguchi R, Saji M, et al.; OCEAN-Mitral Investigators. Clinical impact of baseline frailty status and residual mitral regurgitation after transcatheter edge-to-edge repair: insights from the OCEAN-Mitral registry. J Am Heart Assoc. 2024;13(21):e035109.

3. Shimura T, Yamamoto M, Kano S, Kagase A, Kodama A, Koyama Y, et al.; OCEAN-TAVI Investigators. Impact of the Clinical Frailty Scale on outcomes after transcatheter aortic valve replacement. Circulation. 2017;135(21):2013–24.

4. Proietti M, Romiti GF, Raparelli V, Diemberger I, Boriani G, Dalla Vecchia LA, et al. Frailty prevalence and impact on outcomes in patients with atrial fibrillation: a systematic review and meta-analysis of 1,187,000 patients. Ageing Res Rev. 2022;79:101652.

5. Wilkinson C, Todd O, Clegg A, Gale CP, Hall M. Management of atrial fibrillation for older people with frailty: a systematic review and meta-analysis. Age Ageing. 2019;48(2):196–203.

6. Perera V, Bajorek BV, Matthews S, Hilmer SN. The impact of frailty on the utilization of antithrombotic therapy in older patients with atrial fibrillation. Age Ageing. 2009;38(2):156–62.

7. Volgman AS, Nair G, Lyubarova R, Merchant FM, Mason P, Curtis AB, et al. Management of atrial fibrillation in patients 75 years and older: JACC state-of-the-art review. J Am Coll Cardiol. 2022;79(2):166–79.

8. Gugganig R, Aeschbacher S, Leong DP, Meyre P, Blum S, Coslovsky M, et al.; Swiss-AF Investigators. Frailty to predict unplanned hospitalization, stroke, bleeding, and death in atrial fibrillation. Eur Heart J Qual Care Clin Outcomes. 2021;7(1):42–51.

9. Lefebvre MC, St-Onge M, Glazer-Cavanagh M, Bell L, Kha Nguyen JN, Viet-Quoc Nguyen P, et al. The effect of bleeding risk and frailty status on anticoagulation patterns in octogenarians with atrial fibrillation: The FRAIL-AF Study. Can J Cardiol. 2016;32(2):169–76.

10. Gilhofer TS, Inohara T, Parsa A, Walker M, Uchida N, Tsang M, Saw J. Safety and feasibility of same-day discharge after left atrial appendage closure. Can J Cardiol. 2020;36(6):945–7.

11. Yamamoto M, Otsuka T, Shimura T, Yamaguchi R, Adachi Y, Kagase A, et al.; OCEAN-TAVI Investigators. Incidence, timing, and causes of late bleeding after TAVR in an Asian cohort. JACC Asia. 2022;2(5):622–32.

12. Clegg A, Young J, Iliffe S, Rikkert MO, Rockwood K. Frailty in elderly people. Lancet. 2013;381(9868):752–62.

13. Holmes DR Jr, Doshi SK, Kar S, Price MJ, Sanchez JM, Sievert H, et al. Left atrial appendage closure as an alternative to warfarin for stroke prevention in atrial fibrillation: a patient-level meta-analysis. J Am Coll Cardiol. 2015;65(24):2614–23.

14. Asami M, Naganuma T, Ohno Y, Tani Y, Okamatsu H, Mizutani K, et al.; OCEAN-LAAC Investigators. Initial Japanese multicenter experience and age-related outcomes following left atrial appendage closure: the OCEAN-LAAC registry. JACC Asia. 2023;3(2):272–84.

15. Wang A, Ferro EG, Song Y, Xu J, Sun T, Yeh RW, et al. Frailty in patients undergoing percutaneous left atrial appendage closure. Heart Rhythm. 2022;19(5):814–21.

16. Ong E, Meseguer E, Guidoux C, Lavallée PC, Hobeanu C, Charles H, et al. Prevalence and outcome of potential candidates for left atrial appendage closure after stroke with atrial fibrillation: WATCH-AF Registry. Stroke. 2020;51(8):2355–63.

17. Bibas L, Levi M, Touchette J, Mardigyan V, Bernier M, Essebag V, Afilalo J. Implications of frailty in elderly patients with electrophysiological conditions. JACC Clin Electrophysiol. 2016;2(3):288–94.

18. Holmes DR, Reddy VY, Turi ZG, Doshi SK, Sievert H, Buchbinder M, et al.; PROTECT AF Investigators. Percutaneous closure of the left atrial appendage versus warfarin therapy for prevention of stroke in patients with atrial fibrillation: a randomized non-inferiority trial. Lancet. 2009;374(9689):534–42.

19. Kar S, Doshi SK, Sadhu A, Horton R, Osorio J, Ellis C, et al.; PINNACLE FLX Investigators. Primary outcome evaluation of a next-generation left atrial appendage closure device: results from the PINNACLE FLX trial. Circulation. 2021;143(18):1754–62.

20. Marwick TH, Gillebert TC, Aurigemma G, Chirinos J, Derumeaux G, Galderisi M, et al. Recommendations on the use of echocardiography in adult hypertension: a report from the European Association of Cardiovascular Imaging (EACVI) and the American Society of Echocardiography (ASE). J Am Soc Echocardiogr. 2015;28(7):727–54.

21. Ono S, Kubo S, Maruo T, Kar S, Kadota K. Left atrial appendage size in patients with atrial fibrillation in Japan and the United States. Heart Vessels. 2021;36(2):277–84.

22. Tzikas A, Holmes DR Jr, Gafoor S, Ruiz CE, Blomström-Lundqvist C, Diener HC, et al. Percutaneous left atrial appendage occlusion: the Munich consensus document on definitions, endpoints, and data collection requirements for clinical studies. Europace. 2017;19(1):4–15.

